# Effect of tocilizumab in hospitalized patients with severe pneumonia COVID-19: a cohort study

**DOI:** 10.1101/2020.06.06.20122341

**Authors:** Benjamin Rossi, Lee S. Nguyen, Philippe Zimmermann, Faiza Boucenna, Louis Dubret, Louise Baucher, Helene Guillot, Marie-Anne Bouldouyre, Yves Allenbach, Joe-Elie Salem, Paul Barsoum, Arezki Oufella, Helene Gros

## Abstract

**Background:** Tocilizumab, a drug targeting interleukin-6 administrated in the right timeframe may be beneficial in coronavirus-disease-2019 (COVID-19). We aimed to assess its benefit, drawing from observations in compassionately treated patients.

**Methods:** In a retrospective case-control study, treatment effect (tocilizumab 400mg, single-dose) was assessed using three statistical methods: propensity-score matching, Cox multivariable survival and inverse probability score weighting (IPSW) analyses. Were included all patients hospitalized with COVID-19, who presented severity criteria with SpO2≤96% despite O2-support ≥6L/min for more than 6 hours. Were excluded patients in critical care medicine department and those under invasive mechanical ventilation. Primary outcome was a composite of mortality and ventilation, with a maximum follow-up of 28 days.

**Results:** 246 patients were included (106 treated by tocilizumab). They were 67.6 ±15.3 years-old, with 95 (38.5%) women. Delay between first symptoms and inclusion was 8.4 ±4.5 days. Overall, 105 (42.7%) patients presented the primary outcome, with 71 (28.9%) deaths during the 28-days follow-up. Propensity-score-matched 84 pairs of comparable patients. In the matched cohort (n = 168), tocilizumab was associated with fewer primary outcomes (hazard ratio (HR) = 0.49 (95% confidence interval (95CI) = 0.3–0.81), p-value = 0.005). These results were similar in the overall cohort (n = 246), with Cox multivariable analysis yielding a protective association between tocilizumab and primary outcome (adjusted HR = 0.26 (95CI = 0.135–0.51, p = 0.0001), confirmed by IPSW analysis (p<0.0001). Analyses on mortality with 28-days follow-up yielded similar results.

**Conclusion:** In this retrospective study, tocilizumab single-dose was associated with improved survival without mechanical ventilation in patients with severe COVID-19.

## Introduction

The pandemic of coronavirus, responsible for the severe acute respiratory syndrome (SARS), called Coronavirus Disease (COVID-19) started in the late 2019. It quickly spread worldwide, notably in Europe.(1)

It is responsible for severe pneumonia, resulting in a high rate of transfers to intensive care units (ICU) and in-patients mortality of 5% to 32%.(2)

Severity has been related to an exaggerate immune response, the cytokine release syndrome (CRS), mediated by pro inflammatory cytokines, including interleukin IL-6, IL-12 and tumor necrosis factor α leading to various organs dysfunction including lung, brain and heart.(3) Previously, tocilizumab, an antibody targeting IL-6 receptors proved efficient against CRS.(4) Opposing CRS may decrease further inflammatory pulmonary lesions, i.e. respiratory deterioration requiring mechanical ventilation, transfers to ICU and deaths.(5, 6) Starting on March 23, 2020, we administrated off-label tocilizumab in patients with severe COVID-19 pneumonia as compassionate use. In the present report, we assessed the effect of tocilizumab on mortality and mechanical ventilation in a cohort of patients hospitalized for severe COVID-19 pneumonia. To mitigate selection bias, we performed a triple analysis including propensity-score matching, Cox multivariable and inverse probability score weighting analyses, to compare patients who received tocilizumab, to those who did not.

## Material and method

### Study population

In this observational single-center study, in Robert Ballanger regional hospital, all patients were screened for SARS-coronavirus 2 starting on March 14^th^, 2020. Diagnosis required positive testing with reverse transcription polymerase chain reaction (RT-PCR) or chest CT-scan with typical lesions.(7) Inclusion criteria was severe COVID-19 pneumonia, i.e. pulse oxygen (O2) saturation (SpO2) < 96% despite O2 flow≥6 L/minute delivered by high concentration oxygen mask, for > 6 hours. Were excluded patients under invasive mechanical ventilation and those critical care medicine department. Starting on March 23, 2020, tocilizumab was made available for off-label compassionate use in severe COVID-19 pneumonia in our center. Patients were compared between two group: those who received tocilizumab (a single intravenous injection, 400 mg) and those who did not (control group). This retrospective study was approved by research ethics committee and registered on clinicaltrials.gov (NCT04366206).

### Treatment intervention

Tocilizumab was made available for compassionate use by hospital pharmacists on March 23th, 2020. Choice and indication of treatment depended on attending physician, and information was given to all patients prior to being treated. No criteria was retained to exclude patients from treatment, hence patients without full engagement status (i.e. recused from ICU) were still eligible for treatment.

### Study variables

The primary outcome was a composite of all-cause mortality and invasive mechanical ventilation (i.e. requiring tracheal intubation). Follow-up period was 28 days after inclusion. All data were prospectively collected in electronical medical records, which were then extracted for the purpose of this study. List of available data is presented in *Supplementary Material*. Follow-up was complete for all patients.

### Statistical analyses

Baseline characteristics of patients treated by tocilizumab were compared to that of the control group. In the primary analysis, 1:1 nearest-neighbor propensity-score matching was performed using the following variables: age, sex, smoking status, history of coronary artery disease, stroke, heart failure or peripheral artery disease, hypertension, chronic kidney disease with eGFR less than 60 mL/min/lm73^2^, cancer, long-term corticosteroid treatment, use of antibiotics, of antivirals, of corticosteroids, of baricitinib after admission, SpO2/FiO2 ratio at admission, time between admission and inclusion, and SpO2/FiO2 ratio and CRP at inclusion.(8) A second analysis using a multivariable Cox proportional hazard analysis was performed in the entire cohort, with the following independent covariables: tocilizumab injection, engagement status, age, systolic blood pressure at inclusion, SpO2/FiO2 ratio at admission and SpO2/FiO2 ratio at inclusion. A third analysis using inverse probability score weighting (IPSW) approach was also performed using the entire cohort.(9) Kaplan-Meier curves were used to compare the two groups in the matched cohort. Cox proportional hazards modeling was used to assess the association between tocilizumab and the primary outcome in the overall cohort. Conclusions were drawn only if the three analyses yielded concordant results. Continuous variables were presented as mean ± standard deviation and categorical variables as number (proportion). All analyses were performed using SPSS (IBM, Armonk, USA) and R software.

## Results

In total, 106 patients treated by tocilizumab were included and compared to 140 control patients (flow-chart is presented in **Supplementary Material)**. Overall, 105 (42.7%) patients presented the primary outcome, with 71 (28.9%) deaths during the 28-days follow-up. Baseline characteristics and comparisons are presented in **Table 1**. In the overall cohort (n = 246), patients in the tocilizumab group were younger than those in the control group (64.3 ± 13.0 vs 70.1 ± 16.5 years-old, p< 0.001), more had full engagement status (73 (68.9%) vs 71 (51.4%), p = 0.006), more were treated with antibiotics (106 (100%) vs 134 (95.7%), p = 0.04) and by corticosteroids (43 (40.6%) vs 38 (27.1%), p = 0.27)(see **Supplementary Table 1** for details on overall cohort). In the tocilizumab group, delay between study inclusion and tocilizumab administration was 1.0 ± 1.0 days.

**Table 1.**
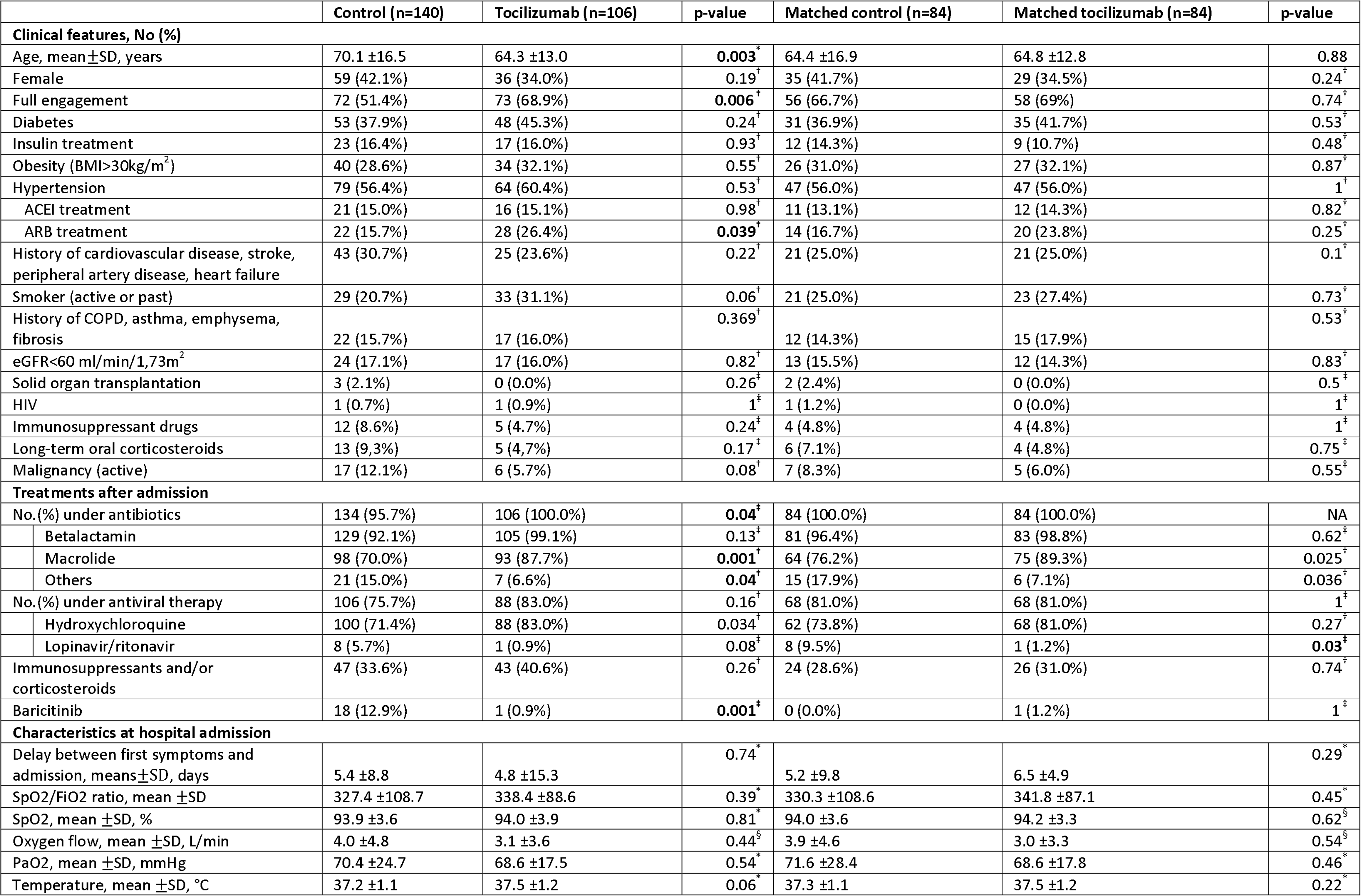
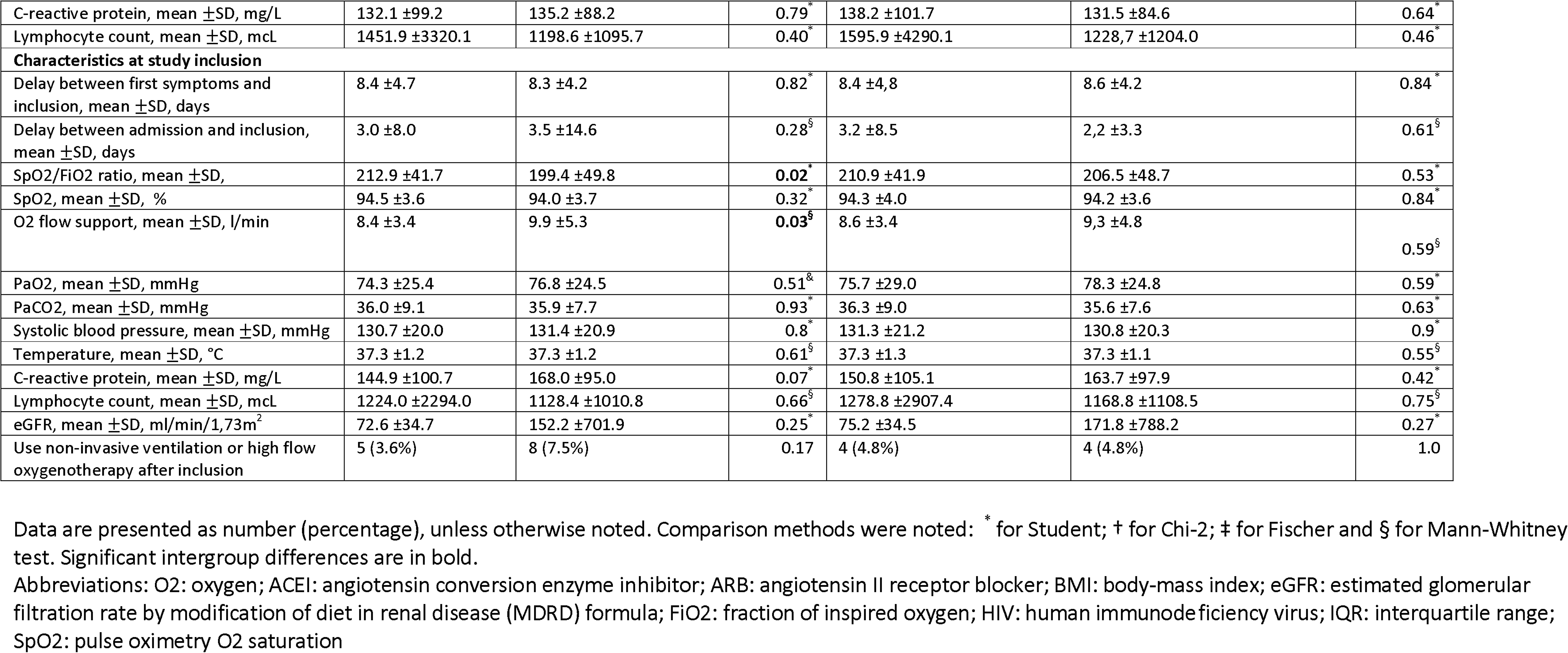
Baseline characteristics, at admission and at inclusion.

### Propensity-score matched cohort

Propensity-score matching yielded 84 pairs of patients (for a total of 168 patients in the matched cohort). There was no significant difference between the two matched groups regarding baseline characteristics (see **Table 1)**. In the matched cohort (n = 168), tocilizumab was associated with fewer events (hazard ratio (HR) = 0.49 (95% confidence interval(95%CI) = 0.30–0.81), p = 0.005)(see **Figure 1)**.

**Figure 1.**
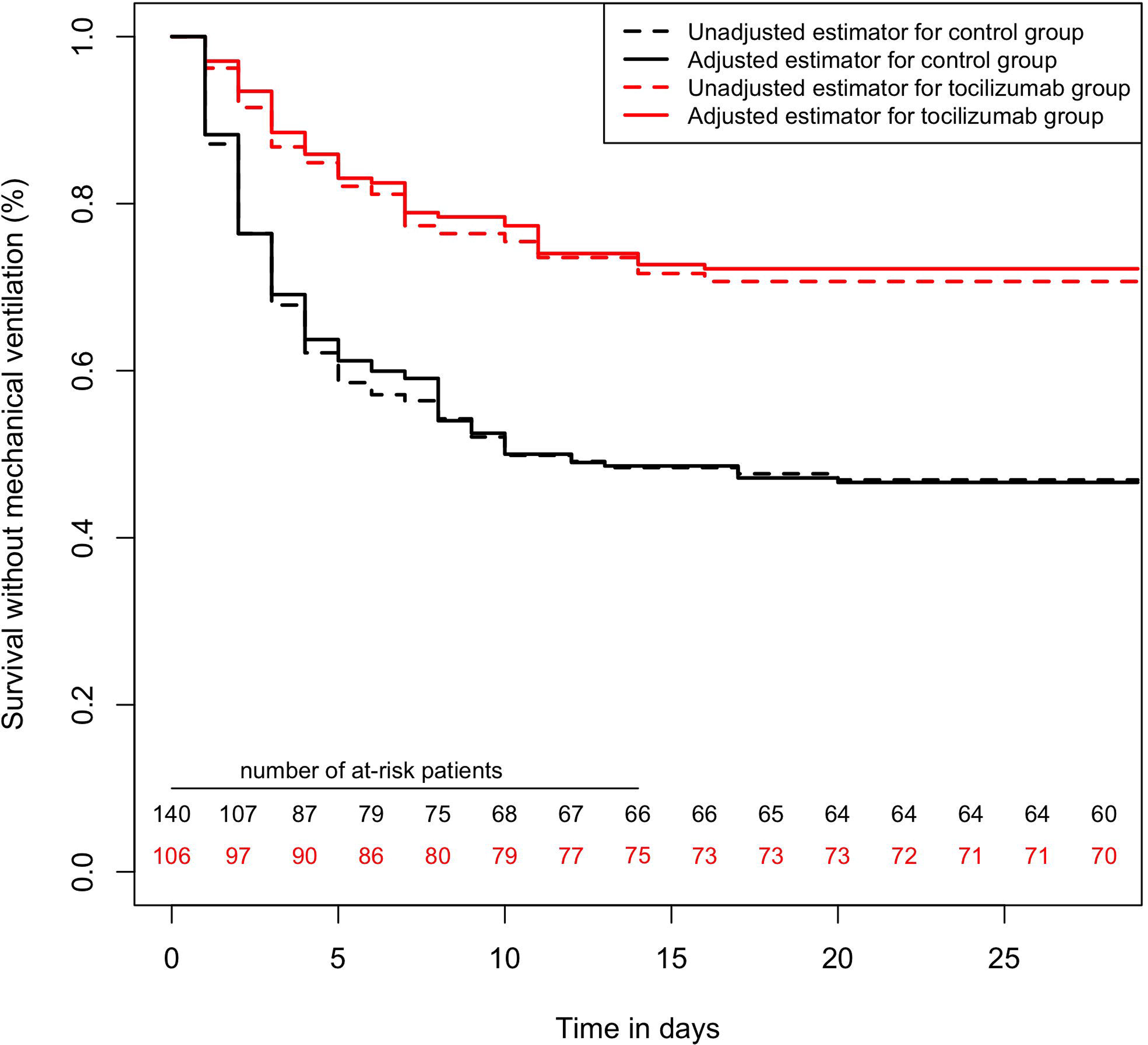
Survival curves, regarding the primary outcome with a 28-days follow-up, comparing tocilizumab and control group. In the matched cohort (n = 168), tocilizumab was associated with fewer events (hazard ratio (HR) = 0.49 (95% confidence interval(95%CI) = 0.30–0.81), p = 0.005). In the overall cohort (n = 246), Cox multivariable survival analysis found tocilizumab independently associated with a lower incidence of the primary outcome (adjusted HR = 0.34 (95%CI = 0.22–0.52), p< 0.0001). Inverse probability score-weighted analysis yielded similar results (p< 0.0001)

### Overall cohort (Cox multivariable and IPSW analyses)

In the overall cohort (n = 246), Cox multivariable survival analysis found tocilizumab independently associated with a lower incidence of the primary outcome (adjusted HR (adj.HR) = 0.34 (95%CI = 0.22–0.52), p< 0.0001). Other variables associated with the primary outcome were: SpO2/FiO2 ratio on the day of inclusion (per 1 unit increase, adj.HR = 0.987 (95%CI = 0.983–0.991), p< 0.0001) and chronic kidney disease (adj.HR = 1.63 (95%CI = 1.03–2.52),p = 0.035). IPSW confirmed the protective association between tocilizumab and primary outcome (p< 0.0001)(see **Figure 1)**.

### All-cause mortality (28-days maximum follow-up)

When considering all-cause mortality with a maximum follow-up of 28 days, in the matched cohort (n = 168), tocilizumab was associated with fewer deaths (HR = 0.42 (95CI= 0.22–0.82), p = 0.008). In the overall cohort (n = 246), Cox multivariable analysis yielded an independent protective association between tocilizumab and mortality (adj.HR = 0.29 (95%CI = 0.17–0.53), p< 0.0001), the other independent variables associated with mortality were full engagement status (adj.HR = 0.11 (95CI= 0.05–0.23), p<0.0001), S/F ratio at inclusion (per 1-unit increase, adj.HR = 0.99 (95CI= 0.985–0.995), p< 0.0001), chronic kidney disease (adj.HR = 2.0 (95CI= 1.22–3.27), p = 0.006) and systolic blood pressure at inclusion (per 1-mmHg increase, adj.HR = 1.016 (95CI= 1.005–1.028), p = 0.006). IPSW analysis was concordant (p< 0.0001)(see **Figure 2)**.

**Figure 2.**
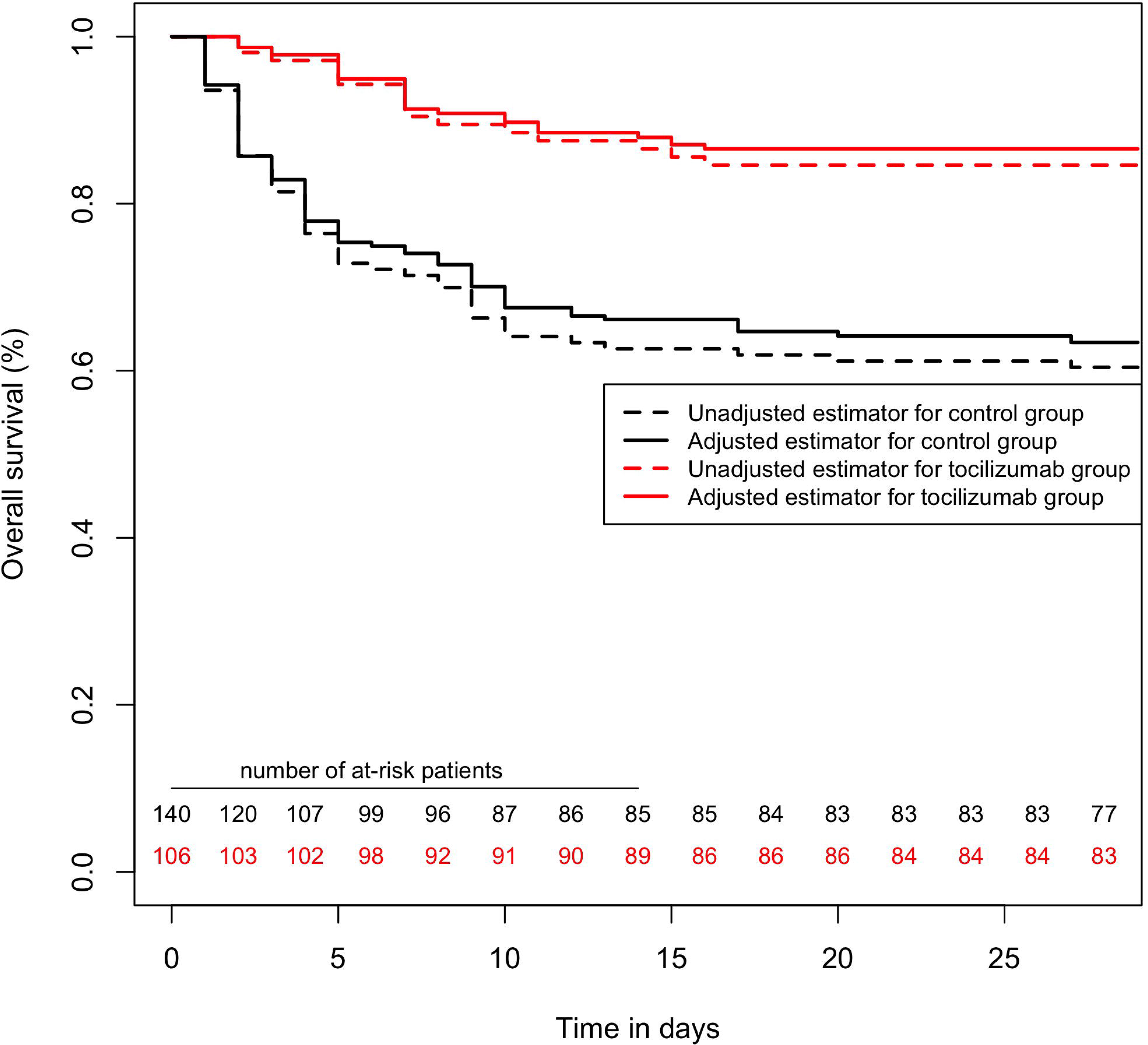
Survival curves, regarding mortality with a 28-days follow-up, comparing tocilizumab and control group. In the matched cohort (n = 168), tocilizumab was associated with fewer deaths (hazard ratio = 0.42 (95CI= 0.22–0.82), p = 0.008). In the overall cohort (n = 246), Cox multivariable analysis yielded an independent protective association between tocilizumab and mortality (adjusted HR = 0.29 (95%CI = 0.17–0.53), p< 0.0001). Inverse probability score-weighted analysis yielded similar results (p< 0.0001).

### Sensitivity analyses

As sensitivity analysis, we focused on the subgroup of patients with full engagement status: 155 patients, including 73 (50.3%) treated with tocilizumab. In these 155 patients, 33 patients presented the primary outcome. Cox multivariable analyses yielded a protective association between tocilizumab and the primary outcome (adj.HR = 0.43 (95CI= 0.22–0.81) p = 0.01); the other variables independently associated with the primary outcome were SpO2/FiO2 ratio at inclusion (per 1-unit increase, adj.HR = 0.985 (95CI= 0.979–0.992) p<0.0001) and temperature at inclusion (per 1°C-increase, adj.HR = 1.39 (950CI= 1.088–1.771), p = 0.008). Similarly, in these 155 patients, 9 died during 28-days follow-up. Cox multivariable analyses yielded a protective association between tocilizumab and the mortality (adj.HR = 0.11 (95CI= 0.01–0.89) p = 0.039).

In a second sensitivity analysis, we excluded patients who presented an outcome during the first 48 hours, to mitigate selection bias (including immortal time bias). This dataset comprised 204 patients, with 97 treated by tocilizumab; 63 patients presented the primary outcome, and 46 died during 28-days follow-up. Using triple statistical method as in the main analysis, tocilizumab was found significantly associated with fewer primary outcomes (Cox multivariable analysis yielded adj.HR = 0.40 (95CI= 0.23–0.70), p = 0.001)(see **Supplementary Figure 1**). Similarly, tocilizumab was found protectively associated with mortality (adj.HR = 0.36 (95CI= 0.18–0.70) p = 0.003)(see **Supplementary Figure** 2)(details of all multivariable models are presented in **Supplementary Material)**.

## Discussion

In this single-center retrospective study in 246 patients, with 106 treated by tocilizumab, we observed a significant association between tocilizumab and fewer deaths and requirement for invasive mechanical ventilation; in patients hospitalized for severe COVID-19 pneumonia, with 28-days follow-up.

The study cohort was similar to that of previously described COVID-19 patients with a median age of 68 years, 27.6% presenting cardiovascular history and 30.1% were obese.(10) Median delay between first symptoms and treatment was 8 days, corresponding to the delay of CRS onset described in SARS-Cov-2.(11)

Attenuating CRS may partly explain the significant decrease in the primary outcome.(5, 6) CRS was related to interleukin accumulation, and a recent trial studying effect of interleukin-1 blockade found significant improvement.(12) Similarly, in our study, clinical improvement observed in patients treated by tocilizumab was akin to that described in a single-center observational study in which 100 patients with Covid-19 were treated with tocilizumab. With 20/100, 17.5% patients who died by day 10, these results are akin to ours (with 28.9% deaths by day 28).(13) Interestingly, in that study, tocilizumab administration protocol mentioned 800 mg twice in 87 patients and 800 mg three times in 13 patients, similar to that of ongoing trials with two injections of up to 800 mg each, within a 3-days period.(14, 15)

In comparison, in our study, dosage of tocilizumab was 400 mg, injected once, following previous reports of improved outcomes chimeric antigen receptor-T-cell-induced CRS with 8 mg/kg dosage.(4) Further confirming our results with the same dosage would improve the availability of this costly biotherapy for which access may become an issue.(16)

We acknowledge several limitations. First, the single-center nature of this study requires external validation, however, it guarantees homogeneity in the care of all patients, in our non-ICU departments dedicated to treat COVID-19 patients, i.e. observed differences are more likely to be due to tocilizumab. Second, although we aimed to mitigate selection bias using three statistical methods, including propensity-score matching, Cox multivariable and IPSW analyses, residual confounders are plausible.(8, 9) Due to comorbidities and lack of beds in the context of Covid-19 pandemic, a significant proportion of patients were labeled as limited, regarding possibilities of being transferred in critical care medicine department, as well as invasive mechanical ventilation. However, these criteria did not alter indications for tocilizumab, as a third was considered limited at admission. All analyses were adjusted for this criterion in multivariable models. Furthermore, we performed additional sensitivity analyses to further mitigate selection bias; analyses which yielded similar results with significant association between tocilizumab and better survival without mechanical ventilation, even focusing on patients with full therapeutic engagement (n = 155). Third, because arterial partial pressure of O2 was not available in all patients, we used SpO2/Fio2 ratio to assess respiratory dysfunction, a validated marker in acute lung injury.(17) Fourth, use of non-invasive ventilation and high-flow oxygenotherapy changed during the study period, following evolving guidelines which advocated against during the first weeks of pandemic to decrease virus aerosol propagation, and were then made more flexible. The relative low proportion of patients who benefited from these treatments and the fact that there was no difference between the two groups regarding this criterion decreases chances of it being a bias although, residual confounding biases may remain. Finally, we did not systematically assay IL-6, which may have proven valuable to identify patients for whom effect was greater.(6)

These results point towards a clinical benefit of tocilizumab, however, they may not replace a full-fledged randomized controlled trial, focusing on dosage adjustment and on patients managed early, so that CRS may be adequately attenuated in patients with Covid-19 evolving towards clinical deterioration.

## Conclusion

In this observational single-center study, in hospitalized patients presenting with severe COVID-19 pneumonia, tocilizumab 400 mg single-dose was associated with lower mortality and need for mechanical ventilation. These results warrant further clinical trials for confirmation and dosage validation.

## Data Availability

data are not available

## Notes

### Competing Interest Statement

The authors have declared no competing interest.

### Clinical Trial

NCT04366206

### Funding Statement

none

### Author Declarations

Regional ethics committee La Sorbonne CER-Sorbonne University (N CER-2020-27) gave us an institutional ethical agreement

